# Bridging the gap from knowledge to action: Implementation of the data to policy (D2P) training program at sub-national levels in Zambia

**DOI:** 10.1101/2023.04.03.23288065

**Authors:** Kutha Banda, Rabson Zimba, Sandra Chilengi-Sakala, Hilda Shakwelele, Olatubosun Akinnola Akinola

## Abstract

Knowledge translation is the synthesis, exchange, and application of knowledge by relevant stakeholders to accelerate the benefits of global and local innovation in strengthening health systems. In Zambia, research evidence is recognized as a critical element for the development of sound policies. This requires deliberate efforts towards generating, harvesting, and utilizing evidence from research and program data to inform decision-making. In response, the National Health Research Authority with support from the Clinton Health Access Initiative adapted a data to policy curriculum for use at sub-national levels and conducted training for 17 healthcare workers. The objectives of the training were to build the capacity of healthcare workers in analyzing research and other data to inform policy and programming as well as to develop six policy briefs for presentation to policymakers.

The curriculum combines epidemiology with economic analysis and modeling to develop informative policy briefs. Sixteen modules were covered and delivered during periodic interactive workshops led by facilitators and mentorship was done in-between sessions. This was done within 6 months from February to August 2022. To assess the participants understanding, Kirkpatrick learning evaluation model was adapted upto level 3; we utilized a pre and posttest method of assessment.

At pre-test, about 71% of the participants scored below 50 percent, while at posttest, all the participants scored above 50%. Six policy briefs were successfully developed covering Sexual Reproductive Maternal Newborn Child Adolescent Health and Nutrition topics. Implementation of this program provided a lot of learnings for programs aimed at improving uptake of evidence into action. One of the key learnings was that conducting economic evaluations and mathematical modelling of proposed policy interventions was critical in informing the decision-makers of the cost and benefits of the interventions. Policy options proposed in the policy brief were largely accepted by key stakeholders and proposed for piloting.

## Introduction

Knowledge Translation (KT) is “the synthesis, exchange, and application of knowledge by relevant stakeholders to accelerate the benefits of global and local innovation in strengthening health systems and improving people’s health.” This entails the scientific process of promoting the uptake of knowledge into practice or strategies used to bridge the gap between research and practice. WHO later defined knowledge translation in public health as “the dynamic interface that links health information and research with policy and practice in an effort to foster evidence-informed policy using different tools (1–3).

Lack of utilization of research results and available knowledge leads to inefficiency and reduction in both quantity and quality of life as there is broad evidence linking translation of evidence to improvement of health care (4). There are many reasons why there is a gap between the production of knowledge and its utilization; some of these include the lack of understanding of policy needs by researchers, research papers being complex in nature and difficult for policymakers to understand and utilize. This, therefore, entails the need for a close relationship among researchers, policymakers, and practitioners (5). To address this gap, the capacity building of both researchers, policymakers and practitioners in knowledge translation is key to achieving desired results in health care (6).

In Zambia, the government has made commendable progress towards achieving its mission of providing quality, cost-effective health services as close to the family as possible by establishing systems that ensure healthcare delivery is in line with this mission. One notable system is evidence-based policy formulation. Research evidence is recognized as a critical element necessary for the development of sound policies, and this is in line with national development strategies. This requires deliberate efforts towards generating, harvesting, and utilizing evidence from research and program data to inform policy formulation, resource allocation and decision-making. To do this, capacities for both the generators of evidence and the users must be built.

In Zambia, knowledge translation has become a leading approach in reducing the gap between research and policy. This has been a technique used since the World Ministerial summit held in Mexico in 2004 (7) which set the goal for knowledge translation as evidence-informed policy and policy-informed research. Subsequent efforts for the African region for knowledge translation system strengthening include 2008’s Bamako call and the research for health 2016-2025 strategy. These have been domesticated at country level and reflected in different national documents including national policies as well as the 8^th^ national development plan, national health, and the national health research strategic plans. This forms a firm ground for knowledge translation and helps in building trust and dialogue among researchers and policymakers.

Identifying the best way to translate research evidence into policy and practice remains an enduring challenge in many health systems across the globe. In an effort to address this challenge in Zambia, the NHRA has been conducting a training program under its knowledge translation function called the Data to Policy (D2P) training with officers in the health sector at the national level. The D2P program is a capacity-building program meant to equip health professionals with the skills to develop evidence-based policy briefs to translate knowledge into an easier-to-understand format for policymakers. The program utilizes evidence from research and existing public health data to package and formulate policy options for policy recommendation and decision-making. It combines epidemiology with economic analysis and modeling to develop informatics, policy and technical briefs.

The D2P program has been implemented at the national level over the past five years since 2017; however, this had not been rolled out to sub-national levels. In order to ensure that this program meets the needs of decision-makers at the sub-national levels and is implemented taking into account the systems at those levels, the NHRA with technical support from the Clinton Health Access Initiative (CHAI) and funding from the Embassy of Sweden in Zambia adapted the national D2P curriculum to develop a curriculum for sub-national levels in 2022. The curriculum aims to ensure that healthcare workers have the skills to address the complexity of knowledge utilization and that knowledge users or policymakers understand the process of integrating research into policy and practice. The curriculum is targeted at frontline officers implementing activities/services in provinces. This model acknowledges Michael Lipsky’s concept of street-level bureaucrats “public service workers who interact directly with citizens in the course of their jobs, and who have substantial discretion in the execution of their work”(8) as key in developing policies.

The D2P curriculum developed for sub-national levels was piloted in Eastern and Southern provinces of Zambia from February to August 2022. This is also in line with the WHO Strategic and Technical Advisory Group of Experts (STAGE) for Maternal, Newborn, Child and Adolescent Health and Nutrition (MNCAHN) recommendations on how to improve knowledge translation in MNCAHN (9). The objectives of the training program were (1) build the capacity of healthcare workers to analyze research and other available data to inform policy and programming of Sexual Reproductive Maternal Newborn Child Adolescent Health and Nutrition (SRMNCAH&N) (2) to develop six policy briefs and present the policy briefs to policymakers. This paper highlights lessons learnt in implementing this program.

## Materials and Methods

The D2P sub-national training program pilot targeted healthcare workers in Eastern and Southern provinces riding on the Peace at the Centre (PeaCe) health program implemented by the Ministry of Health in the provinces alongside CHAI with support from the Embassy of Sweden. This program covers a total of 16 modules (10) adapted from the national level D2P program. The modules were delivered during periodic interactive workshops led by trained facilitators.

### Modules

The curriculum and modules adoption was done in consultation with representation from CHAI, NHRA, the Zambia National Public Health Institute, the University of Zambia-School of public health, the Tropical Diseases Research Centre and two officers from the pilot provinces. The modules in this curriculum include:

1. Zambia knowledge translation story. This provided an overview of the country’s efforts of strengthening knowledge translation through integrating it in the different laws, regulations and guidelines developed to this effect.
2. Introduction to data to policy. Gives an overview of content covered in the program.
3. Policy and policy briefs. The module gives an introduction to public health policies, types of policies and how policies are made.
4. Literature search and data sources. Provides a guide to the different types of data, where it can be found and how to search for it.
5. Problem statement and root cause analysis. Provides a guide on how to frame a problem and root cause analysis using tools such as fishbone diagram in order to arrive at critical causes.
6. Epidemiologic measures in policy briefs. This modules covers several measures of diseases such as population attributable risk, statistical significance and how this data can be used to tell a story.
7. Identification of policy options. This module covers policy analysis by assessing the public health impact, budgetary impact, economic impact and feasibility as well as assessing the validity of the evidence used.
8. Stakeholder identification and mapping. The module covers several steps of stakeholder analysis including stakeholder identification, mapping, interviews and analysis and provides different tools for this.
9. Framing an economic evaluation. This covers different economic evaluation methods for partial and full analysis.
10. Assessing policy options health impacts. The module covers how to assess the health impact of the proposed policy interventions by building models. Several tools can be used, including the decision tree in Microsoft excel, which is free of charge.
11. Cost analysis. This module focuses on calculating costs and the different types of costs to account for in the different interventions.
12. Economic evaluation; Cost-effectiveness analysis and budget impact analysis. This module guides on the different types of economic evaluation and different formulas used for calculating these as well as budget impact analysis.
13. Sensitivity analysis. This teaches the participants how to conduct sensitivity analysis.
14. Effective writing. This module guides the participants how to use different skills to simplify the analysis for effective communication to different audiences.
15. Making and writing recommendations. This module gives tip to learners on how to make actionable recommendations.
16. Advocacy strategy and communication. The module provides a guide on what policy advocacy is and key strategies that can be used.

### Course format

We delivered the course within 6 months (from February to August 2022) through virtual and in-person sessions. The project, took in seventeen participants recommended by the Provincial Health Offices (PHOs) – 8 from Eastern and 9 from Southern province. The participants were put in 6 groups of 2-3 members to work as a team on developing the policy briefs based on an agreed topic of primary importance in their province. Each team had 2 or 3 mentors assigned to them: 1 or 2 focusing on the public health and epidemiological aspects, and 1 focusing on costing and economic modelling of recommendations and assumptions of the policy briefs.

Three week-long main learning and writing sessions/workshops were held in-person covering 4-5 modules of the curriculum each. A fourth in-person workshop was also held for 5 days to focus on writing the policy briefs and preparation of presentations for the policy forum. These were interactive in nature to allow diverse thinking on each topic. Two other workshops lasting 5-8 days (half days) were held virtually for mentors to work with mentees on developing the policy briefs. Furthermore, weekly check-in sessions usually lasting an hour were held between mentors and mentees in each group, these sessions were useful for each group to discuss the assigned deliverables.

Additional to the learning and writing sessions, the D2P program employs the integrated Knowledge Translation (iKT) technique where stakeholders are engaged throughout the policy briefs writing process The stakeholders engaged included technical experts and members of Technical Working Groups (TWGs) on the specific topics from national to sub-national levels.

At the beginning of the training, participants were made aware of the course objectives and deliverables, and to assess their understanding of the key concepts of the course, Kirkpatrick learning evaluation model (11) was adapted up to level 3. The model has four levels including:

1. Reaction: This level evaluates how the learners felt, and their personal reactions to the training or learning experience
2. Learning: This is the measurement of the increase in knowledge or intellectual capability from before to after the learning experience
3. Behavior: This is the extent to which the learners applied the learning and changed their behavior, and this can be immediately and several months after the training.
4. Results: This is the effect on the environment resulting from the improved performance of the learner - it is the acid test

The Kirkpatrick model was adapted and used up to level 3 to inform program improvement. Level 1 and 2 of the evaluations were based on the learning environment, these levels focused on the D2P course delivery, mentorship and understanding of the content. Level 3 of the evaluation focused on the performance of the learners in terms of their ability to analyze, synthesize data and produce policy briefs. To analyze this data, we utilized a pre and posttest method of assessment using survey questionnaire Survey CTO. This data was analyzed using STATA software package.

## Results and Discussion

In this section, we share the social demographic characteristics of the participants and their pre and post-training scores. We further share lessons learnt in implementing the program and summarize key outcomes.

Table 1 shows that about 41% of the participants were aged between 30 and49: of these, 65% of them were male. Fifty nine percent of the participants had master’s degrees and 41% were medical doctors by training. The participants’ designations included managers, program officers, planners and information officers with the majority of them (24%) being District Health Directors (DHDs)

**Table 1:** Social-demographic characteristics of participants

| **Demographic characteristics** |  |  |
| --- | --- | --- |
| **Characteristics** |  |  |
|  | **Freq.**<br>**(N=17)** | **Percent** |
| **Age group** |  |  |
| 18-29 | 1 | 6 |
| 30-39 | 7 | 41 |
| 40-49 | 6 | 35 |
| 50-59 | 3 | 18 |
| **Sex** |  |  |
| Female | 6 | 35 |
| Male | 11 | 65 |
| **Highest education attained** |  |  |
| Diploma | 1 | 6 |
| Degree | 4 | 24 |
| Masters | 10 | 59 |
| PhD | 2 | 12 |
| **Training background** |  |  |
| Demography | 1 | 6 |
| Environmental Health | 1 | 6 |
| Medical Doctor | 7 | 41 |
| Monitoring and Evaluation | 1 | 6 |
| Nursing/Midwifery | 4 | 24 |
| Nutritionist | 2 | 12 |
| Planner | 1 | 6 |
| **Designation** |  |  |
| Clinical care specialist | 1 | 6 |
| District Health Director | 4 | 24 |
| Nursing Officer | 3 | 18 |
| Nutritionist | 2 | 12 |
| Planner | 1 | 6 |
| Provincial Quality Assurance and Quality Improvement Program Officer | 1 | 6 |
| Senior Health Information Officer | 2 | 12 |
| Senior Health Promotion Officer | 1 | 6 |
| Senior Medical Superintendent | 2 | 12 |

### Objective 1: To build the capacity of healthcare workers to analyze research and other available data to inform policy and programming of Sexual Reproductive Maternal Newborn Child Adolescent Health and Nutrition (SRMNCAHN)

We administered a pre and post-test to the participants to gauge their knowledge of the different concepts in the curriculum. This included questions on policy analysis, epidemiology and economic analysis. We found that at pre-test, about 71% of the participants scored below 50 percent, while at post-test, all the participants scored above 50% (table 2).

**Table 2:** Overall performance of participants

| Knowledge level |  |  |
| --- | --- | --- |
| Characteristics |  |  |
|  | Freq. (N=17) | Percent |
| **Aggregate percentage score pre-test** |  |  |
| <50 | 12 | 71 |
| 50+ | 5 | 29 |
| **Aggregate score post-test** |  |  |
| 50+ | 17 | 100 |

### Objective 2: To develop six policy briefs and present the policy briefs to policy makers

Six policy briefs were developed during the mentorship entitled:

1. It Isn’t Just a Little Rash! Protect a child from measles by improving uptake of the second measles vaccine dose through mandatory birth registration and vaccination tracking in Southern province.
2. How Many Newborn Babies Can We Save By Booking Antenatal Care Early? Introduce free early community pregnancy detection in Southern province.
3. Iron for a Strong, Smart and Healthy Child! Reducing the high prevalence of anaemia in children aged 6-23 months in Southern Province, Zambia.
4. Save Adolescent Girls’ Lives! Enhanced contraceptive uptake key to reducing maternal mortality in adolescents.
5. Born Alive to Thrive! Ending preventable stillbirths in Eastern Province
6. Trauma-Informed Care! A solution to post intimate partner violence depression management in Eastern Province, Zambia

The policy briefs were presented at provincial policy forums with several stakeholders. These were further used to engage policymakers including 10 parliamentarians at the 10^th^ Zambia Heath Research Conference. The policy forum provided an opportunity for discussion of the proposed interventions as compared to the status quo and the recommendation from this meeting was to lobby for resources to pilot the recommended policy options in the respective provinces.

### Lessons learned

Implementation of this program provided a lot of learnings for programs aimed and improving uptake of evidence into action. Some of these lessons include:

1. The importance of involving the implementers in the process of developing policies cannot be overemphasized. Having the program implementers with understanding of the practical challenges they face to be the ones to analyze data and develop policy recommendations was key to acceptance of results and henceforth implementation.
2. Involving Implementers that generate the routine data to develop the policy briefs made them appreciate their role and importance of generating quality data for it to be informative.
3. Developing policies at sub-national levels acknowledging the decision making powers at those levels is important in contextualizing interventions to those a particular environment
4. Engagement of stakeholders including community members in the process from problem identification, identification of policy options and evaluation of the policy options was critical for buy-in and ownership of results/recommendations
5. Conducting economic evaluations and mathematical modelling of proposed interventions was critical in informing the decision-makers of the cost and benefits of the proposed interventions

### Challenges

#### Time constraint due to competing priorities

The time commitment of the participants was a challenge, given that they all run extremely busy offices. The time constraint was mitigated by setting time expectation from the beginning as the program is highly involving for detailed literature review and data analysis as well as economic analysis. Further, an orientation of the program was provided to the supervisors of the officers to get their buy-in and continuous engagement was done to ensure availability of the participants amidst other competing priorities.

#### Access to data

Another challenge encountered was access to some articles which were not open access particularly those which presented results of effective interventions in similar settings. In some cases, journal articles which seemed to have data on effective interventions and their cost was not open access and this inhibited access for the participants. Another challenge noted was the limited context applicable data on effective interventions.

## Conclusion

There are several challenges faced in health care. These include challenges in the improvement of health care systems and reducing the risk of adverse events, failure to optimally use evidence generated in research greatly contributes to these challenges especially in developing countries like Zambia. Effective translation of research evidence into policy and practice is important for improving health outcomes and reducing health inequities, and this essentially addresses the “know-do” gap (12); one of the barriers to the utilization of research results is that researchers misunderstand the needs of decision-makers and consequently, decision-makers may not use existing research evidence in their decision-making. The D2P program provided an opportunity for researchers, practitioners and decision-makers to engage in an effort to close the “know-do” gap.

Furthermore, to steer clear of spending billions annually in public and private sectors of biomedical, clinical, and health services research, it is important that healthcare professional training and continuing professional development in KT programs such as D2P is enhanced (13). This program was largely accepted by key stakeholders including parliamentarians who recommended having regular dialogues of this nature to achieve the government’s mission of providing quality, cost-effective health services as close to the family as possible.

## Data Availability

The data used in this analysis from the program participants is available and uploaded with this submission. However, the actual policy briefs developed are not uploaded with this submission as those will be submitted for publication too

## Acknowledgments

We would like to acknowledge the Centre for Disease Control and Prevention Foundation (CDCF) through the data for health initiative, vital strategies and the Bloomberg data for health initiative for developing the original D2P curriculum which was adapted for the sub-national curriculum. We also thank the National Health Research Authority for managing the D2P program. Further thanks go to the mentors who provided their time to teach and mentors the participants and the participants from Eastern and Southern province of Zambia for taking this course and engaging with key stakeholders. Lastly, we would like to thank the Embassy of Sweden in Zambia for finding the program and making this achievement possible.

